# Two-Step Machine Learning to Diagnose and Predict Involvement of Lungs in COVID-19 and Pneumonia using CT Radiomics

**DOI:** 10.1101/2022.06.15.22276090

**Authors:** Pegah Moradi Khaniabadi, Yassine Bouchareb, Humoud Al-Dhuhli, Isaac Shiri, Faiza Al-Kindi, Bita Moradi Khaniabadi, Habib Zaidi, Arman Rahmim

## Abstract

**Objective:** We aimed to develop a two-step machine learning (ML) based model to diagnose and predict involvement of lungs in COVID-19 and non COVID-19 pneumonia patients using CT chest radiomic features.

**Methods:** Three hundred CT scans (3-classes: 100 COVID-19, 100 pneumonia, and 100 healthy subjects) were enrolled in this study. Diagnostic task included 3-class classification. For severity prediction, two radiologists scored involvement of lungs in COVID-19 and pneumonia scans based on percentage of involvement in all 5 lobes. Datasets were classified into mild (0-25%), moderate (26-50%), and severe (>50%). Whole lungs were segmented utilizing deep learning-based segmentation method. Altogether, 107 features including shape, first-order histogram, second and high order texture features were extracted. For both tasks, datasets were randomly divided into 90% training sets (70% and 30% for training and validation, respectively) and 10% test sets. Pearson correlation coefficient (PCC≥90%) was performed to exclude highly correlated features. Subsequently, different feature selection algorithms (Correlation attribute evaluation, Information gain attribute, Wrapper Subset selection algorithm, Relief method, and Correlation-based feature selection) were assessed. The most pertinent features were finally selected using voting method based on the evaluation of all algorithms. Several ML-based supervised algorithms were utilized, namely Naïve Bays, Support Vector Machine, Bagging, Random Forest, K-nearest neighbors, Decision Tree and Ensemble Meta voting. The synthetic minority oversampling technique (SMOTE) was used to balance the three classes in training sets. The optimal model was first selected based on precision, recall and area-under-curve (AUC) by randomizing the training/validation sets 20 times, followed by testing using the test set. To ensure the repeatability of the results, the entire process was repeated 50 times.

**Results:** Nine pertinent features (2 shape, 1 first-order, and 6 second-order features) were obtained after feature selection for both phases. In diagnostic task, the performance of 3-class classification using Random Forest was 0.909±0.026, 0.907±0.056, 0.902±0.044, 0.939±0.031, and 0.982±0.010 for precision, recall, F1-score, accuracy, and AUC, respectively. The severity prediction task using Random Forest achieved 0.868±0.123 precision, 0.865±0.121 recall, 0.853±0.139 F1-score, 0.934±0.024 accuracy, and 0.969±0.022 AUC.

**Conclusion:** The two-phase ML-based model accurately classified COVID-19 and pneumonia patients using CT radiomics, and adequately predicted severity of lungs involvement. This 2-steps model showed great potential in assessing COVID-19 CT images towards improved management of patients.

## 1. Introduction

The COVID-19 pandemic has infected nearly 500 million individuals across the globe, with over 6 million death cases reported in the world [1]. Meticulous COVID-19 screening allows for early and accurate diagnosis, minimizing the impact on healthcare systems while also saving lives and limiting the disease’s spread [2, 3]. Specifically, chest X-ray and CT scans are the first accurate diagnostic tools for determining the types of COVID-19 [4].

Radiomics analysis has great potential for precision medicine as it uses data mining to create a correlation between clinical and biological findings [5, 6]. Explicit radiomics features such as shape, statistics and texture are derived from medical images, resulting in accurate and non-invasive COVID-19 biomarkers that potentially influence clinical decision making [3, 7]. Artificial intelligence (AI) based methods, including Machine Learning (ML), have been used in several COVID-19 research efforts [8, 9]. In fact, ML-based models using CT-based radiomics, increasingly utilized for diagnosis, severity prediction and prognosis of COVID-19 disease are straightforward and time efficient.

As examples, Tang *et al*. [10] assessed CT radiomics of 176 COVID-19 patients and COVID-19 was detected with higher accuracy using Random Forest (RF) model. According to Liu *et al*. [11], the decision curve evaluation demonstrated the clinical applicability of the COVID-19 radiomics model for detection by means of deriving a radiomics signature compared to C-RADS. Fang *et al*. [12] studied a total of 75 CT images (46 COVID-19 and 29 pneumonia). They extracted 77 explicit radiomics from the total lesions in the lungs. Multiple cross-validation was utilized to choose the primary characteristics following SVM as classifier to validate/test the AI-based model.

Recently, Shiri *et al*. [7] combined radiological data, clinical data, and CT radiomics features to develop a novel prognostic models for prediction (alive, death) on a total of 152 COVID-19 patients. Based on a multivariant analysis, model obtained 0.95 AUC, 0.89 accuracy, 0.88 sensitivity and 0.89 specificity. Elsewhere, Cai *et al*. [13] aimed to explore how COVID-19 pneumonia severity evaluation and clinical outcome prediction in COVID-19 patients were modified by CT measurements. The severity of disease was classified into three levels: moderate, severe, and critical. To identify the severity of the illness, they constructed random forest and regression model. They reported AUCs of RF classifiers in the classification of moderate vs. (severe + critical) and severe vs. critical were 0.93 of both classifications.

In a large cohort multicenter studies by CT radiomics features extracted from Lung regions and machine learning from 14339 and 26307 patients, prognostic [14] and diagnostic [3] models were developed, respectively. In a most recent study [15], COVID-19 severity scoring was performed using CT radiomics features and multinomial multiclass machine learning models. More than thousand COVID-19 patient’s data from four different class of severity were enrolled. Bagging random forest (BRF) and multivariate adaptive regression (MAR) splines coupled with multinomial logistic regression were implemented. They reported precision of 0.86 and 0.82, recall of 0.84 and 0.79, accuracy of 0.93 and 0.92 and AUC of 0.82 and 0.82 for BRF and MAR algorithms respectively.

Most recent studies have focused on either employing explicit radiomics to diagnose and differentiate COVID-19 pneumonia from other viral pneumonias or using ML techniques to stratify disease severity. However, to our knowledge, no research has been published on the validity of CT radiomics towards ultimate decision-making in the management of COVID-19 and pneumonia patients simultaneously in terms of diagnosis and severity prediction of clinical outcomes. We thus aimed to develop and deploy a two-phase ML radiomics signature approach. The model was developed based on radiomics features extracted exclusively from the whole lungs of CT images for differential diagnosis of COVID-19 and pneumonia to adequately evaluate their severity. Early diagnosis of COVID-19 and pneumonia etiology, particularly in patients with a highly suspected, may help clinicians in implementing appropriate patient management plans and reducing triage time during hospital admissions. In addition, severity prediction of COVID-19 and pneumonia, may help radiologists in making rapid diagnosis, that is particularly important when the healthcare system is overloaded. In what follows, we elaborate on our methods, followed by results, discussion, and conclusion.

## 2. Materials and Methods

### 2.1 Data collection and segmentation

Imaging data from 300 CT scans (100 COVID-19 cases, 100 Pneumonia cases, and 100 Healthy ones) were collected from the Sultan Qaboos University Hospital (SQUH) and Royal Hospital (ROYH), Muscat, Oman. The SQUH and ROYH medical research ethical committees both approved this retrospective study (MREC#1254-REF. NO. SQU-EC/121/20). COVID-19 cases met the following criteria: (a) RT-PCR positive, (b) non-contrast CT on chest CT. Aside the COVID-19 group, ground truth of pneumonia cases was reviewed based on the radiologists’ reports on CT images. CT Chest scans were performed on the Siemens Somatom definition flash 128 slices. Tube voltage, rotation time and pitch were 120 kVp, 0.6s and 1.55 respectively. The reconstructed series matrix size was 512×512 pixels with 1 mm slice thickness. The scans were performed at maximum inspiration.

For segmenting whole lungs, DICOM CT images were segmented utilizing automated deep learning (DL) based segmentation for lungs and COVID-19 pneumonia infectious lesions (COLI-Net) which we have previously developed and extensively evaluated [16]. In this study we only employed the whole lung segmentation. Figure 1 illustrates CT images and segmentations of Healthy, COVID-19 and pneumonia cases. All segmentations were evaluated by human observer to be sure about its accuracy and in case of mis-segmentation, segmentation was manually edited.

**Figure 1.**
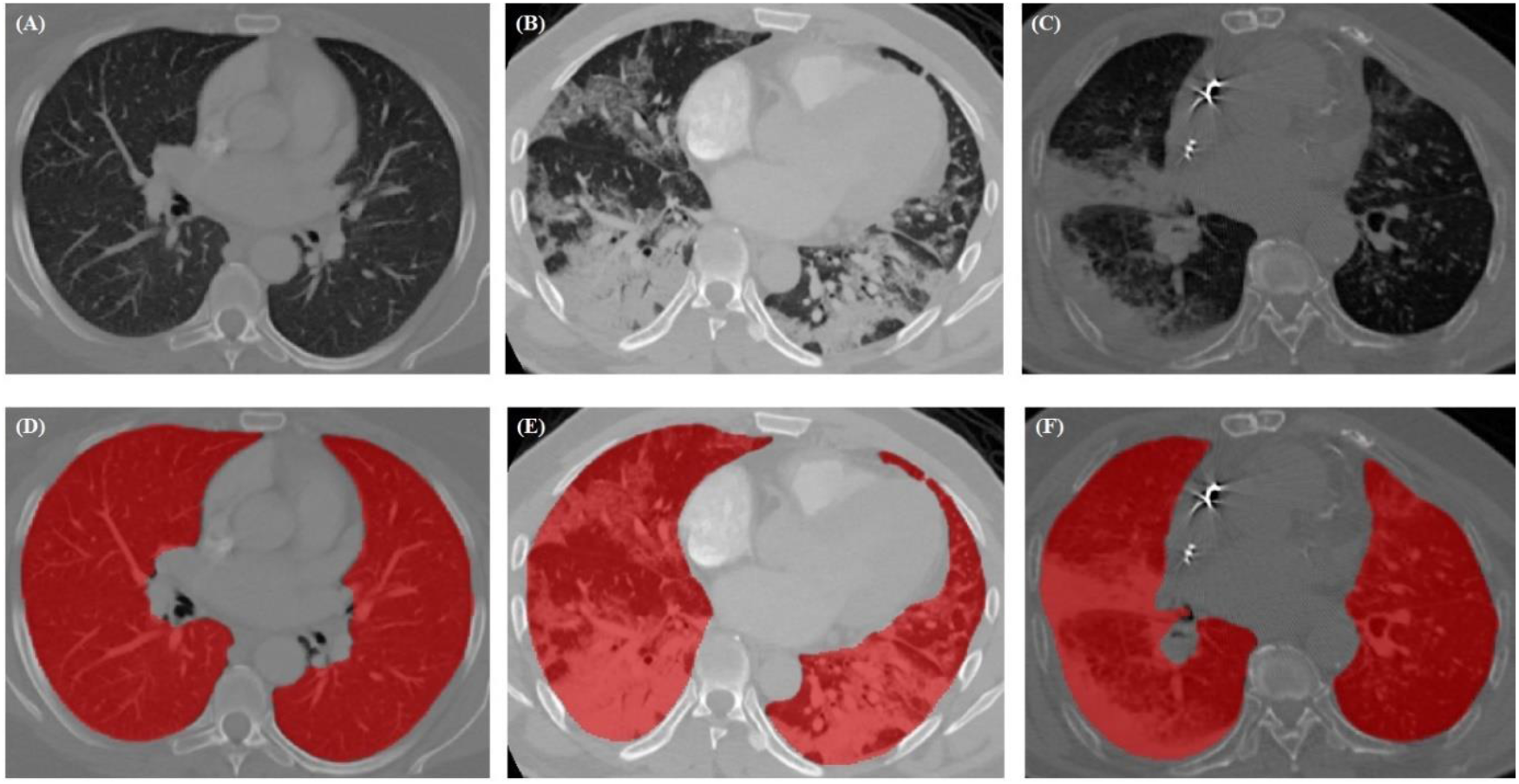
Segmentation of Healthy (*left*); COVID-19 (*middle*); Pneumonia (*right*) utilizing our DL-based lungs segmentation method (COLI-Net).

### 2.2 Severity scoring

The standardized score CO-RADS was utilized to assess the severity of infection of the whole lungs, enabling to analyze COVID-19 cases based on CT image findings [17]. Thus, for severity prediction, two experienced radiologists scored in consensus pulmonary involvement of COVID-19 RT-PCR positive and Pneumonia scans by percentage of involvement within all 5 lobes. The percentage of each lobe (right upper, right middle, right lower, left upper, and left lower) affected by the pulmonary opacities such as ground-glass, mixed ground-glass and consolidation, consolidation, organizing pneumonia, etc were included. Scores were recorded as (a) 0: 0% involvement; (b) score 1: < 5% involvement; (c) score 2: 5%–25% involvement; (d) score 3: 26%–50% involvement; (e) score 4: 51%–75% involvement; and (f) score 5: > 75% lobar involvement). Total lungs involvement (labeled as subjective severity score) was concluded by adding the scores from all lobes (minimum score 0; maximum score 25). Finally all cases were classified based on the severity score of all lobes into two groups (extensive: total score ≥ 15; non-extensive: total score < 15) [18]. For developing the prediction model, the infection involvement of both lungs was classified into mild (0-25%), moderate (26-50%), and severe (>50%) is illustrated in the figure 2.

**Figure 2.**
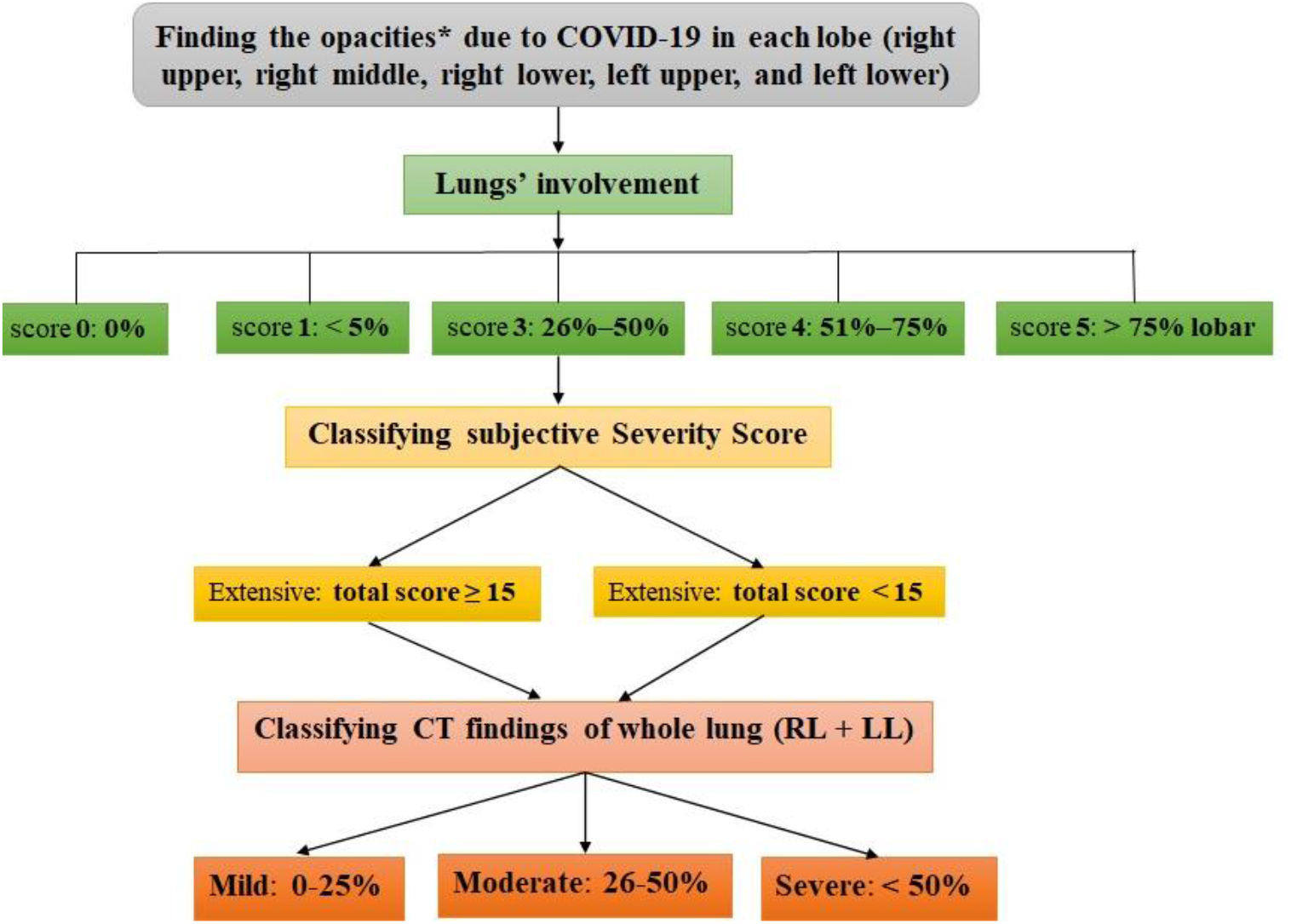
A flowchart of severity scoring method. (*\*OPACITIES: Ground-glass, Mixed ground-glass and Consolidation, Consolidation, Reverse halo sign with ground-glass opacity surrounded by consolidation, Nodular, or Ground-glass with Crazy paving appearance. RL: Right Lung, LL: Left Lung*)

### 2.3 Feature extraction and dimensionality reduction

The 107 features namely shape, first-order histogram, second and high order texture features, were generated using the Pyradiomics library [19]. All images were resized to isotropic voxel size 2×2×2 mm^3^ and image intensity discretized by 64-gray level binning, followed by feature extraction.

These features were divided into three categories: (i) intensity, (ii) shape, and (iii) texture. Lung intensity-based features, defined using first order statistics of the intensity histogram, quantified the tissue density of the right and left lungs on chest CT images. Shape features describe the 3D geometric properties of the lungs, whereas textural features quantified the infected region heterogeneity. Textural features were computed by analyzing the spatial distribution of voxel intensities in thirteen directions. Explicit features were computed from gray level co-occurrence (GLCM), run length matrices (GLRLM), Gray Level Dependence Matrix (GLDM), Gray Level Size Zone Matrix (GLSZM), and Neighborhood Gray-Tone Difference Matrix (NGTDM). It worth noting that the Pyradiomics image analysis software (PIAS) is part of the Image Biomarker Standardization Initiative (IBSI) [20].

To reduce the highly correlated features, prior to feature selection, Spearman correlation coefficient (PCC ≥ 90%) was carried out. Subsequently, for selection of the most relevant features, 5 different feature selection algorithms including Correlation attribute evaluation, information gain attribute, wrapper subset feature selection, Relief method, and correlation-based feature selection using WEKA (version 3.8.2) were deployed. Unlike other studies [21, 22], the most pertinent features were finally selected using voting method by evaluating the performances of algorithms. We have chosen features that were frequent among more than 4 algorithms in this performance assessment stage.

### 2.4 Model Derivation and Validation

#### I. Diagnostic phase

Datasets were randomly divided into 90% training sets and 10% test sets. The training set was used to select the best models (consistent with best practice guidelines) [23], and the test sets were used to report on the performance of the selected classifiers. The training sets was again split into 70% training and 30% validation sets; where the training set in this step of AI pipeline was used for learning and the validation set was used for giving an estimate of the model performance. Test sets were untouched during validation.

We considered several ML-based supervised algorithms namely Naïve Bays, Support Vector Machine, Bagging, Random Forest, K-nearest Neighbors, Decision Tree and Ensemble Meta Voting, as utilized through the WEKA toolkit. To select the optimal model based on precision, recall and AUC, we randomized training/validation sets 20 times. Splitting into training and validation set was performed 20 times to reduce variability in estimation of the model performance. The optimal model and Ensemble Meta Voting were then tested using the test set. We computed precision, recall, F1-score, accuracy, and AUC. To ensure the repeatability of the results, assessment using the test set, the entire operation was repeated 50 times.

The framework in figure 3 was first developed from scratch step by step for the diagnosis task (classification as: COVID-19, Pneumonia or Healthy) and then using the same selected features (same 8 features mentioned above) we considered the 3 classes (mild, moderate, and severe) for the prediction step.

**Figure 3.**
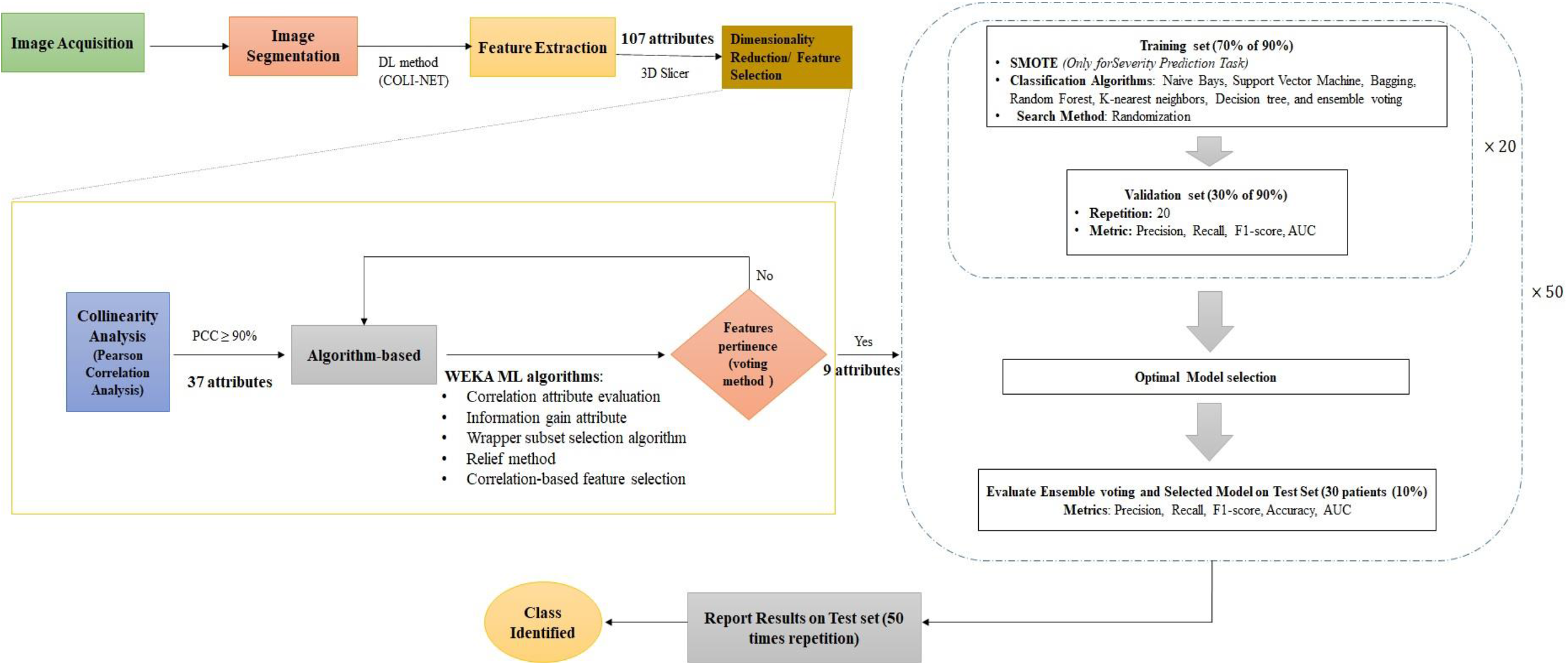
An AI workflow in this study. Segmentation was performed using DL method while other steps were based on ML methods

#### II. Prediction of severity phase

For severity prediction, two experienced radiologists scored involvement of lungs in COVID-19 and Pneumonia scans based on percentage of involvement in all 5 lobes. In severity prediction phase, previous datasets were classified into mild (0-25%), moderate (26-50%), and severe (>50%). In this second prediction phase the 300 datasets were randomly divided into 90% training set and 10% test sets. The training set was again split into 70% training and 30% validation sets, and test sets were untouched during validation. Prior to training, the synthetic minority oversampling technique (SMOTE) was used to balance the three classes (mild, moderate, severe). Note that for the diagnosis phase, the entire dataset was classified into 3 equal classes (healthy, COVID-19 and pneumonia), and therefore, SMOTE algorithm was only applied for the predication phase. Training and validation were performed as described in the diagnostic phase. Briefly, by using pre-selected radiomics, we considered several ML-based algorithms (as outlined in previous section). To select the optimal model based on precision, recall and AUC, we randomized training/validation sets 20 times. Splitting into training and validation set was performed 20 times to reduce variability in estimation of the model performance. The optimal model and ensemble meta voting tested using the test set. The model performance parameters (precision, recall, F1-score, accuracy, and AUC) were reposted. To ensure the repeatability of the results, assessment using the test set, the entire operation was repeated 50 times.

## 3. Results

To minimize overfitting and build a robust radiomics signature, dimensionally reduction (correlation-based feature selection method) reduced the number of features from 107 to 37. Further reduction to 9 features was achieved using the algorithm-based feature. The 9 most pertinent features include: shape features (2), first-order features (1), and second-order features (6), respectively. Spearman correlation, narrowed down the number of features to 37 features by removing redundant features (see Figure 4) [24]. To distinguish between informative features from redundant/noisy and irrelevant features 5 different supervised features selection algorithms were employed [25]. Then the results were compared utilizing voting method. In this step, 28 more features were eliminated. Hence, a set of 9 non-redundant and relevant features (shape flatness; shape least length; first order kurtosis; GLCM cluster prominence; GLCM cluster shade; GLCM ImC1; GLDM small dependence high gray level emphasis; GLRLM run length nonuniformity; and GLSZM small area emphasis) were employed in constructing the two-phase ML-based model.

**Figure 4.**
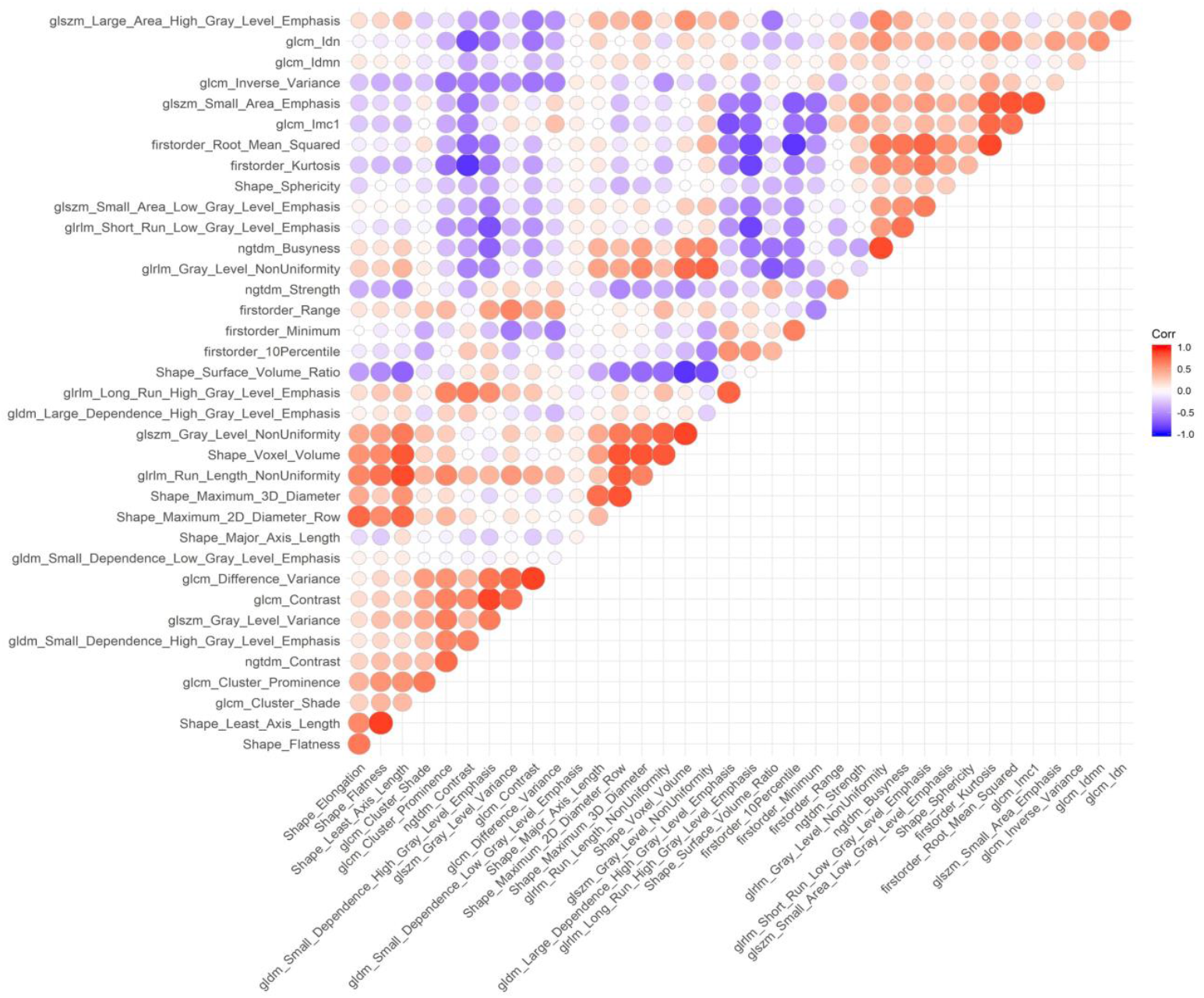
Correlation matrix between the extracted features.

Table 1 and Figure 5 summarize the results of optimum model selection with 20 times cross validation for each class (healthy, COVID-19, pneumonia). Random forest in comparison to the other classifiers achieved to best classification with precision = 0.922 (0.017), recall = 0.922 (0.057) and AUC = 0.0979 (0.014).

**Table 1.** Performance of several classifiers (mean(SD)) for the selection of the optimal model.

| Classifier | Class | Precision | Recall | AUC |
| --- | --- | --- | --- | --- |
| **Random Forest** | Healthy | 0.907(0.052) | 0.977(0.026) | 0.990(0.008) |
|  | COVID-19 | 0.942(0.049) | 0.926(0.060) | 0.984(0.017) |
|  | Pneumonia | 0.919(0.054) | 0.863(0.072) | 0.963(0.022) |
|  | Macro-Ave | **0.922(0.017)** | **0.922(0.057)** | **0.979(0.014)** |
| **Meta voting** | Healthy | 0.891(0.063) | 0.992(0.019) | 0.966(0.017) |
|  | COVID-19 | 0.927(0.049) | 0.935(0.054) | 0.944(0.041) |
|  | Pneumonia | 0.932(0.056) | 0.826(0.076) | 0.890(0.038) |
|  | Macro-Ave | 0.916(0.022) | 0.917(0.084) | 0.933(0.039) |
| **Naïve Bays** | Healthy | 0.824(0.047) | 0.998(0.005) | 0.968(0.018) |
|  | COVID-19 | 0.760(0.076) | 0.902(0.065) | 0.969(0.021) |
|  | Pneumonia | 0.866(0.079) | 0.562(0.098) | 0.874(0.042) |
|  | Macro-Ave | 0.816(0.053) | 0.82(0.229) | 0.937(0.054) |
| **Support Vector Machine** | Healthy | 0.815(0.050) | 0.964(0.051) | 0.945(0.027) |
|  | COVID-19 | 0.821(0.071) | 0.855(0.037) | 0.907(0.043) |
|  | Pneumonia | 0.823(0.075) | 0.656(0.080) | 0.774(0.047) |
|  | Macro-Ave | 0.819(0.004) | 0.825(0.156) | 0.875(0.089) |
| **K-nearest-neighbor** | Healthy | 0.931(0.047) | 0.927(0.058) | 0.943(0.027) |
|  | COVID-19 | 0.896(0.058) | 0.923(0.064) | 0.932(0.040) |
|  | Pneumonia | 0.871(0.074) | 0.841(0.08) | 0.877(0.048) |
|  | Macro-Ave | 0.899(0.030) | 0.897(0.048) | 0.917(0.035) |
| **Bagging** | Healthy | 0.831(0.066) | 0.962(0.046) | 0.979(0.016) |
|  | COVID-19 | 0.880(0.083) | 0.864(0.073) | 0.969(0.023) |
|  | Pneumonia | 0.838(0.087) | 0.734(0.103) | 0.922(0.035) |
|  | Macro-Ave | 0.849(0.026) | 0.853(0.114) | 0.956(0.030) |
| **Decision Tree** | Healthy | 0.874(0.061) | 0.909(0.076) | 0.942(0.035) |
|  | COVID-19 | 0.889(0.065) | 0.897(0.097) | 0.924(0.044) |
|  | Pneumonia | 0.840(0.095) | 0.789(0.091) | 0.867(0.056) |
|  | Macro-Ave | 0.867(0.025) | 0.865(0.066) | 0.911(0.039) |

**Figure 5.**
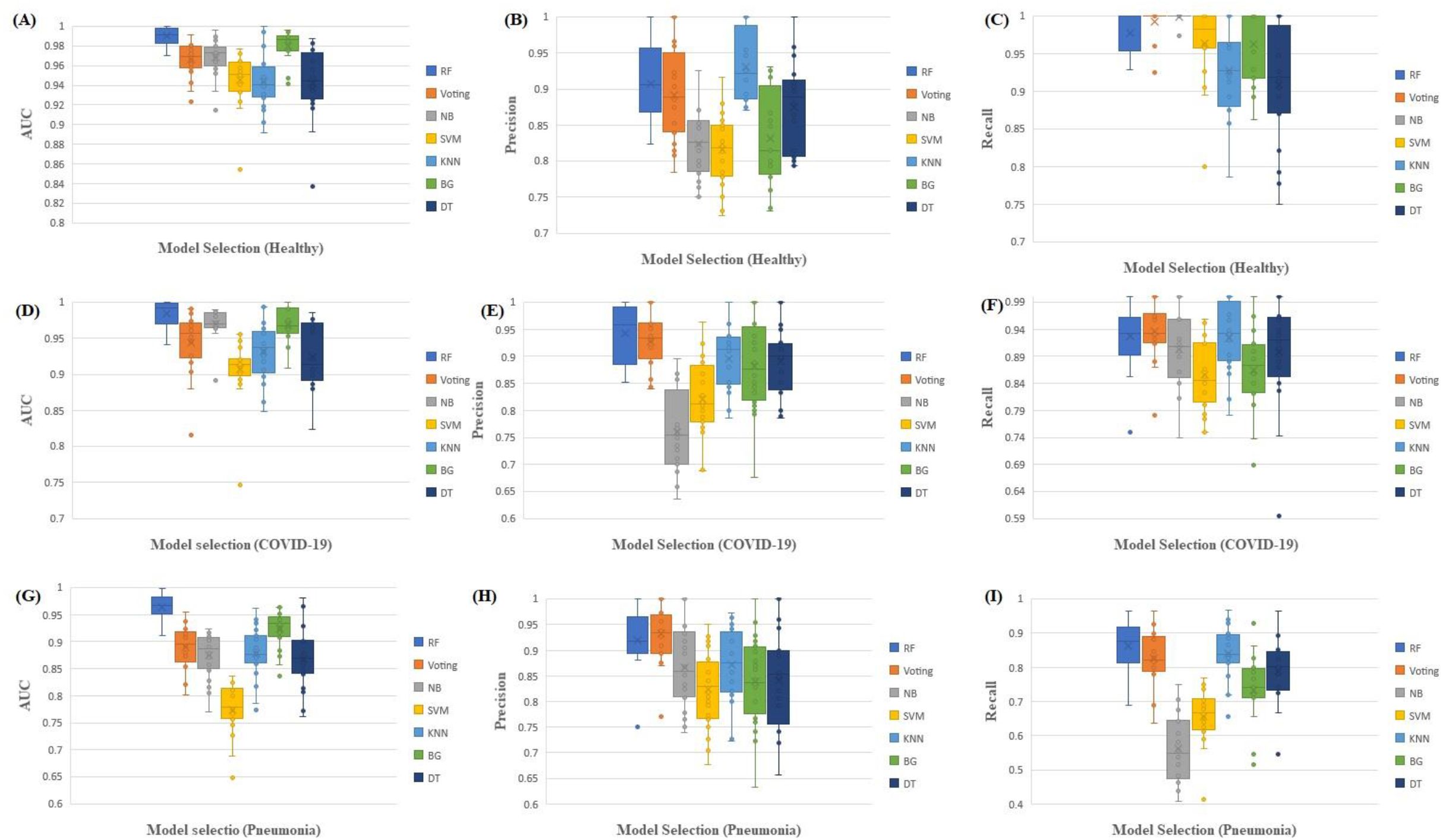
Boxplots of the AUC, Precision, and recall for different classier algorithms and classifications; healthy (A,B,C), COVID-19 (D,E,F), pneumonia (G,H,I).

For the diagnostic task, the performance of the 3-class classification (healthy, COVID-19, pneumonia) using Random Forest and Meta Voting algorithm was assessed by computing precision, recall, F1-score, accuracy, and AUC. Table 2 shows the performance of the Random Forest and Meta Voting methods on test sets. The macro average scores for 3-class classification are provided to indicate the overall performance across the different classes of validation and test sets. From the validation utilizing Random Forest and Meta Voting, precision was 0.922±0.017 and 0.916±0.022, recall was 0.922±0.057 and 0.917±0.084 and AUC was 0.979±0.014 and 0.933±0.039, respectively. Moreover, based on the 3×3 confusion matrix, for RF and meta voting, the precision was 0.909±0.026 and 0.894±0.011, recall was 0.907±0.056 and 0.897±0.078, and AUC was 0.982±0.010 and 0.922±0.043, respectively in testing.

**Table 2.** Classification (healthy, COVID-19, pneumonia) performance (mean (SD)) indices by random forest and meta voting.

|  | Classifier | Class | Precision | Recall | F1-score | Accuracy | AUC |
| --- | --- | --- | --- | --- | --- | --- | --- |
| Test | RF | Healthy | 0.934(0.090) | 0.969(0.077) | 0.951(0.065) | 0.970(0.032) | 0.993(0.011) |
|  |  | COVID-19 | 0.913(0.010) | 0.893(0.116) | 0.894(0.079) | 0.940(0.039) | 0.984(0.020) |
|  |  | Pneumonia | 0.881(0.105) | 0.859(0.111) | 0.863(0.078) | 0.908(0.046) | 0.969(0.024) |
|  |  | Macro-Ave | **0.909(0.026)** | **0.907(0.056)** | **0.902(0.044)** | **0.939(0.031)** | **0.982(0.010)** |
|  | Meta voting | Healthy | 0.908(0.104) | 0.969(0.073) | 0.934(0.077) | 0.962(0.033) | 0.964(0.044) |
|  |  | COVID-19 | 0.888(0.097) | 0.910(0.086) | 0.893(0.05) | 0.929(0.042) | 0.927(0.043) |
|  |  | Pneumonia | 0.887(0.096) | 0.813(0.119) | 0.844(0.077) | 0.894(0.049) | 0.877(0.061) |
|  |  | Macro-Ave | **0.894(0.011)** | **0.897(0.078)** | **0.89(0.045)** | **0.928(0.034)** | **0.922(0.043)** |
| Validation | RF | Healthy | 0.907(0.052) | 0.977(0.026) | 0.939(0.026) | 0.962(0.020) | 0.990(0.008) |
|  |  | COVID-19 | 0.942(0.049) | 0.926(0.060) | 0.94(0.037) | 0.956(0.025) | 0.984(0.017) |
|  |  | Pneumonia | 0.919(0.054) | 0.863(0.072) | 0.887(0.042) | 0.919(0.024) | 0.963(0.022) |
|  |  | Macro-Ave | **0.922(0.017)** | **0.922(0.057)** | **0.922(0.030)** | **0.945(0.023)** | **0.979(0.014)** |
|  | Meta voting | Healthy | 0.891(0.063) | 0.992(0.019) | 0.938(0.031) | 0.959(0.023) | 0.966(0.017) |
|  |  | COVID-19 | 0.927(0.049) | 0.935(0.054) | 0.930(0.037) | 0.956(0.023) | 0.944(0.041) |
|  |  | Pneumonia | 0.932(0.056) | 0.826(0.076) | 0.874(0.047) | 0.914(0.024) | 0.890(0.038) |
|  |  | Macro-Ave | **0.916(0.022)** | **0.917(0.084)** | **0.914(0.034)** | **0.943(0.025)** | **0.933(0.039)** |

Similar to the diagnostic phase, the best model performance was achieved using the Random Forest algorithm. Furthermore, the performance of Random Forest and meta voting algorithms for severity prediction based on most pertinent radiomics was evaluated. Table 3 summarizes the performance of random forest and meta voting for severity prediction. It was also possible to classify CT scans with mild, moderate, and severe using random forest algorithm with the precision of 0.868±0.123, recall of 0.865±0.121, and AUC of 0.969±0.022. Moreover, performance of severity prediction model using meta voting algorithm showed precision of 0.86±0.123), recall of 0.849±0.149, and AUC of 0.895±0.072.

**Table 3.** Classification (mild, moderate, severe) performance (mean (SD)) indices by random forest and meta voting.

|  | Classifier | Class | Precision | Recall | F1-score | Accuracy | AUC |
| --- | --- | --- | --- | --- | --- | --- | --- |
| Test | RF | Mild | 0.958(0.058) | 0.960(0.054) | 0.957(0.039) | 0.962(0.035) | 0.987(0.018) |
|  |  | Moderate | 0.728(0.230) | 0.729(0.233) | 0.694(0.197) | 0.915(0.050) | 0.944(0.055) |
|  |  | Severe | 0.920(0.075) | 0.908(0.081) | 0.908(0.072) | 0.927(0.049) | 0.976(0.029) |
|  |  | Macro-Ave | **0.868(0.123)** | **0.865(0.121)** | **0.853(0.139)** | **0.934(0.024)** | **0.969(0.022)** |
|  | Meta voting | Mild | 0.959(0.059) | 0.965(0.053) | 0.959(0.037) | 0.965(0.032) | 0.959(0.064) |
|  |  | Moderate | 0.722(0.242) | 0.680(0.244) | 0.663(0.202) | 0.910(0.056) | 0.816(0.122) |
|  |  | Severe | 0.899(0.092) | 0.902(0.098) | 0.894(0.075) | 0.915(0.054) | 0.911(0.062) |
|  |  | Macro-Ave | **0.86(0.123)** | **0.849(0.149)** | **0.838(0.155)** | **0.93(0.030)** | **0.895(0.072)** |
| Validation | RF | Mild | 0.938(0.037) | 0.965(0.031) | 0.951(0.22) | 0.961(0.017) | 0.986(0.014) |
|  |  | Moderate | 0.905(0.065) | 0.902(0.050) | 0.901(0.034) | 0.946(0.019) | 0.981(0.019) |
|  |  | Severe | 0.910(0.042) | 0.880(0.057) | 0.893(0.033) | 0.928(0.021) | 0.975(0.010) |
|  |  | Macro-Ave | **0.917(0.017)** | **0.915(0.044)** | **0.915(0.031)** | **0.945(0.016)** | **0.98(0.005)** |
|  | Meta voting | Mild | 0.937(0.043) | 0.946(0.032) | 0.941(0.022) | 0.955(0.016) | 0.950(0.025) |
|  |  | Moderate | 0.878(0.070) | 0.918(0.049) | 0.850(0.199) | 0.941(0.018) | 0.935(0.023) |
|  |  | Severe | 0.924(0.048) | 0.876(0.060) | 0.897(0.038) | 0.932(0.025) | 0.919(0.031) |
|  |  | Macro-Ave | **0.913(0.031)** | **0.913(0.035)** | **0.896(0.045)** | **0.942(0.011)** | **0.934(0.015)** |

On the other hand, the performance validation metrics for the validation sets were precision of 0.917±0.017, recall of 0.915±0.044, and AUC of 0.98±0.005. Moreover, performance of severity prediction model using meta voting algorithm showed precision of 0.913±0.031, recall of 0.913±0.035, and AUC (of 0.934±0.015. The model accuracy and validation show how well model is generalizing.

## 4. Discussion

CT imaging scans have been widely used in COVID-19 studies. High quality 3D CT images have been utilized for COVID-19 identification and patient management. Since early days of the pandemic, numerous ML-based studies were focused on different tasks including diagnosing, prognostic, severity prediction utilizing binary or multiple classifications, using CT scans as illustrated in Table 4 [21, 26-31]. Radiomics has attracted significant attention since early pandemic studies as a fully non-invasive tool for an in-depth quantitative analysis of CT images. The CT radiomics framework uses sophisticated quantitative features in CT images (“radiomics features”) to describe types of lesions and tissue patterns [3]. These characteristics are grouped into two categories: semantic and agnostic characteristics. Agnostic features (e.g., textural features) apply creative mathematical processes in a high-throughput fashion that may not be possible to be detected by the human eye. Moreover, semantic features are used to define morphologic properties of lesions such as shape, size, location, etc. On chest CT scans, we extracted radiomics features associated with healthy, COVID-19, and pneumonia, which provides a noninvasive method to identify radiomic patterns of COVID-19 and pneumonia, differentiate COVID-19 from pneumonia and healthy, and predict COVID-19 and pneumonia severity.

**Table 4:**
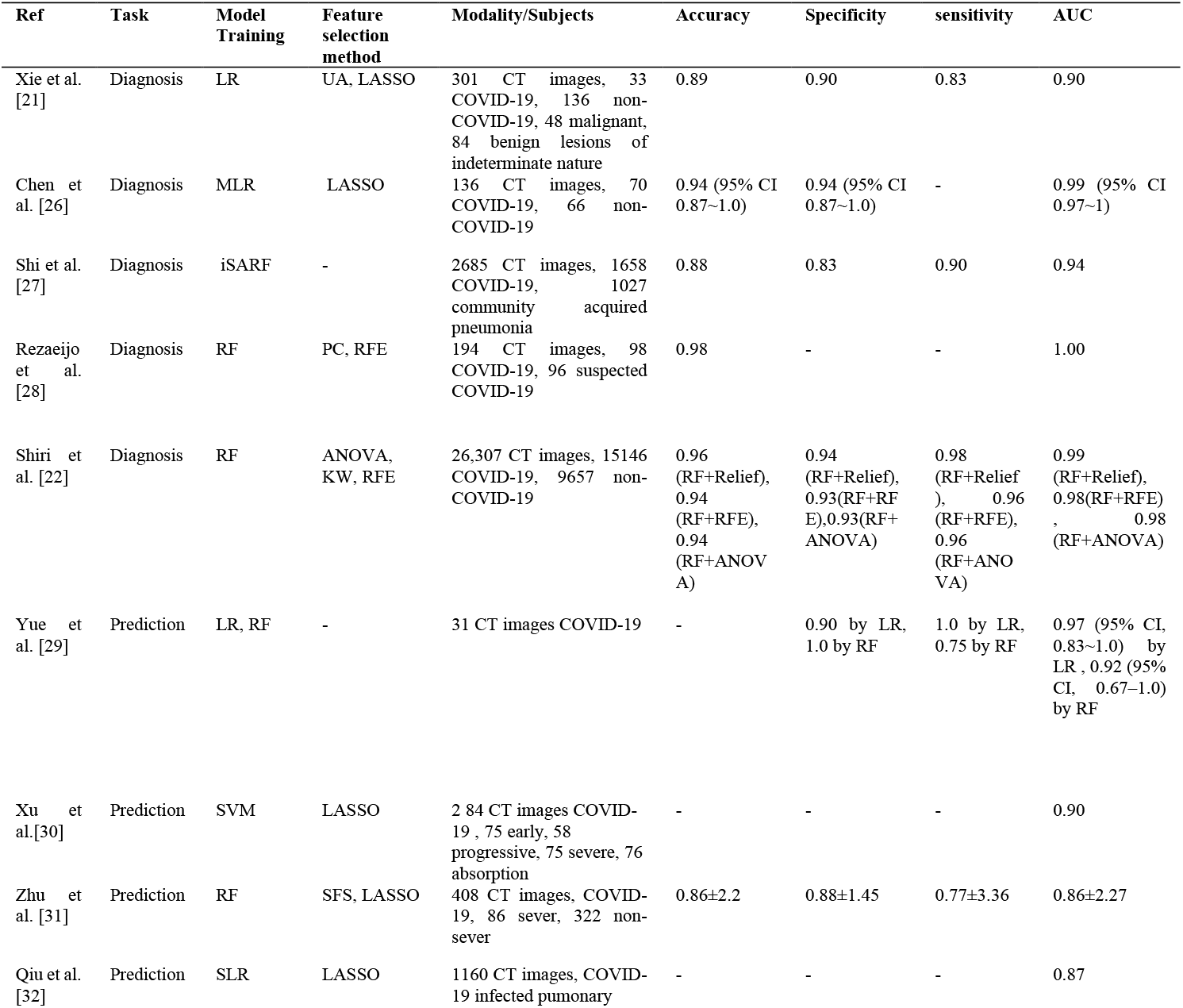
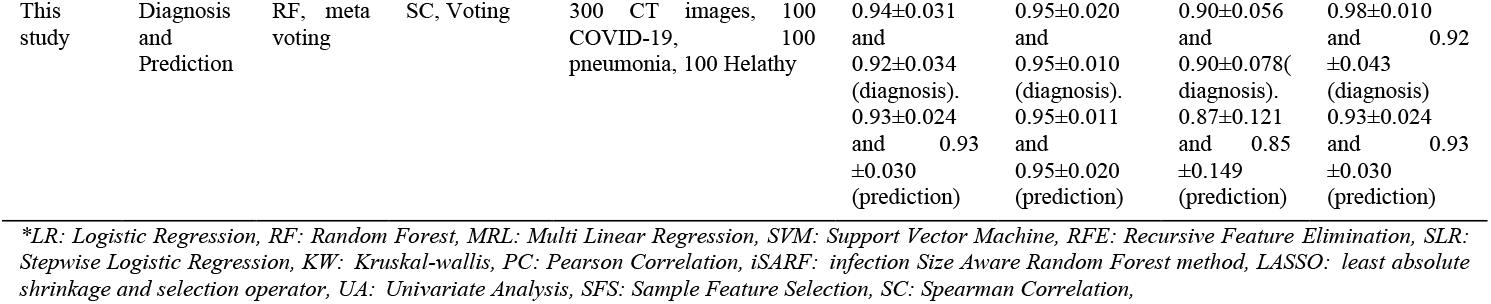
ML-based models used in COVID-19.

In AI modeling for diagnosis, Xie *et al*. [21] used 301 CT images from three hospitals (33 COVID-19 and 268 pneumonia). The ground glass opacity (GGO) lesion was used to assess their CT radiomic model that provided an AUC of 0.90. LASSO and univariant analysis were deployed to select features, and logistic regression was used to classify them. The model assisted in distinguishing COVID-19 from other etiologies in a population confounded by GGO alterations. Similarly, Chen *et al*. [26] evaluated the diagnosis ability of ML-based model 70 COVID-19 and 66 non-COVID-19 from five centers with as AUC of 0.99 (95% CI 0.966∼1.000). Besides GGO, they included (p = 0.032) consolidation in the lung periphery as well as the lesion size.

Another study by Shi *et al*. [27] comprised 1,658 COVID-19 patients and 1,027 community-acquired pneumonia patients who received thin-section CT. For distinguishing COVID-19 from pneumonia, an infection size-aware random forest technique (iSARF) was proposed. With an AUC of 0.94, experimental data demonstrate that the proposed technique performs best when explicit features were being used. Additional studies on 734 patients using thick slice images showed the potential of a generalizable model. Huang *et al*. [33] analyzed the diagnostic value of CT-based signs combined with radiomics features to discriminate COVID-19 from other viral pneumonia. A total of 181 CT scans (89 COVID-19, and 92 non-COVID-19) were utilized. In the training and the testing cohort, the model achieved an AUC of 0.90 and 0.87, respectively

Rezaeijo *et al*. [28] suggested a ML-based model for diagnosis task utilizing data from lesion segmentation of CT images (98 COVID-19 and 96 suspected COVID-19). A total of 755 radiomics characteristics were extracted, the most relevant features obtained using Pearson Correlation and Recursive Feature Elimination. The model was also trained using random forest, decision tree, and K-nearest neighbor classifiers. For the feature selection technique RFE and the RF classifier, they obtained AUC=1.00. A Recent study by Shiri *et al*. [22] indicated without the use of any other diagnostic test, CT-based radiomics derived from lung, paired with ML method, can enable highly effective identification of COVID-19.

In AI modelling for prediction, Xu *et al*. [30] assessed the CT-based model to the predictive ability of the support vector machine for COVID-19 severity in four groups: early, progressive, severe, and absorption. Macro-average of four class-classifications yielded AUCs of 0.97 and 0.9, respectively, for the training and test sets. Zhu *et al*. [31] suggested a novel COVID-19 severity prediction ML-based model that combined the disease progression and the conversion time. Experimental analysis was performed in two hospitals with 408 CT images. Results indicated that ML-based model achieved an AUC of 0.86±2.27. Yue *et al*. [29] enrolled 52 individuals from five different centers in a retrospective analysis. In the first four centers, CT radiomics models were based on logistic regression and random forest that assisted using features from pneumonia lesions. On a lung lobe and patient level, the prediction performance was examined in the fifth center (test dataset). The CT radiomics provided an AUC of 0.97 (95 percent CI, 0.83–1.0) for short-term admission (less than 10 days) in the hospital and an AUC of 0.92 (95 percent CI, 0.67–1.0) for long-term admission (> 10 days), according to LR and RF, respectively.

Elsewhere, Pourhomayoun and Shakibi [34] used 7 different ML algorithms (logistic regression, support vector machine, decision tree, neural networks, random forest, and K-nearest neighbor) to predicted mortality risk of COVID-19 by using CT images. With an overall accuracy of 0.90, the Neural Network technique performed significantly better in predicting the death rate. Zhou *et al*. [35] employed a ML-based model to predict the progression of sickness severity. They used a genetic algorithm (GA) and support vector machine algorithm for feature selection and prediction, respectively. Recently, Wungu *et al*. [36] employed supervised learning to investigate the link between a variety of cardiac indicators and COVID-19 severity and/or mortality, and they concluded that high CK-MB, PCT, NT-proBNP, BNP, and d-dimer levels could be predictive markers of COVID-19 severity.

In this study we utilized 9 explicit radiomics features using voting method to develop the 2 phase ML-based model for diagnosis and severity prediction of COVID-19 and pneumonia. Generally, a radiomics features can distinguish minor changes in the region of interest, that are not be visible to a naked eye, by a computer algorithm. Shape features illustrate properties of the size and shape of the images, which was found to be effective in the classification/severity prediction of COVID-19 and pneumonia infections. Indication of shape feature dimensions was suggested to be investigated during each phase of COVID-19 [30, 37, 38].

First-order statistics represent the distribution of voxel intensities within the region of interest that significantly related to the pixel values on the CT scan through basic metrics. The first-order features mainly indicate the internal texture of lung regions [39]. Kurtosis is one of first-order features, that is used to compute of the “peakedness” of the distribution of the values. Luís *et al*. [40] confirmed that COVID-19 induces consolidation and ground glass opacification, resulting in lower kurtosis values and flatter peak.

A gray level co-occurrence matrix (GLCM) provides the arrangement of voxel pairs to determine texture such as homogeneity (a reflection of the uniformity) of the distribution of voxels. Moreover, the gray run length matrix (GLRLM) provides the size of the uniform run per each gray level [41]. Recently, Hongmei *et al*. [42] illustrated that uniformity was less for COVID-19 than lesions in the non-COVID-19 class, with a greater range in irregular texture for the COVID-19 class. They indicated a more heterogeneous lung texture possibly on the grounds of diversity in airspace disease phenotypes such as crazy paving on visual inspection.

Moura *et al*. [40] observed more heterogeneity of GLRLM in the upper left region of COVID-19 class which may be due to consolidation that tended to diffuse. Pizzi *et al*. [37] predicted that the increasing homogeneity in COVID-19 might correlate with the degree of inflammatory infiltrate in the early stage of diffuse alveolar damage. In the severe phase, however, ground glass opacity develops in density and heterogeneity, forming a crazy paving pattern. The Gray Level Zone Length Matrix (GLZLM) gives parameters about the uniform zone size of each gray level in 3D or 2D region of interest. The gray-level nonuniformity of the COVID-19 group tended to be larger in compared to those of non-COVID-19 ones [41]. Moreover, GLSZM quantifies grey-level zones in an image as the number of connected voxels that share the same grey-level intensity.

For the first task (diagnosis), the validation of our 2 phase ML-based model utilizing RF and meta voting algorithms achieved AUCs of 0.979±0.014 and 0.933±0.039, respectively. For testing, RF and meta voting achieved AUCs of 0.982±0.010 and 0.922±0.043, respectively. Despite, for the second task (severity prediction) validation of our model achieved AUC of 0.969±0.022 using RF algorithm and AUC of 0.895±0.072 for meta voting. For testing, AUCs are 0.98±0.005 and 0.934±0.015 for RF and meta voting, respectively.The gap between the training and validation accuracy indicates the amount of overfitting [43]. In this study the percentage difference between accuracy of validation and testing for diagnosis task are 1.1% and 0.2% for RF and meta voting, respectively. Moreover, the gap between accuracy of validation and testing in severity prediction are 0.3% and 0.4% for RF and meta voting, respectively.

Despite addressing bias and limitations in establishing a generalizable model, some aspects should be considered in evaluating the results: (i) our proposed model is based on a medium sample size, however for full generalizability, multi-centric/large datasets are required. As a result, using accurate data from the open source COVID-19 data repository could increase the model’s accuracy; (ii) our model does not include clinical, demographic and laboratory data. However, previous studies showed significant correlation of CT radiomics with these findings [7, 18, 26, 36]; (ii) we enrolled in-patient COVID-19 positive and pneumonia. Therefore, the moderate group was significantly lower than the mild and severe groups. To solve this issue, SMOTE algorithm was employed on training set only to obtain reproducible and repeatable performance; (iv) we did not include the impact of lesion biologic heterogeneity of lungs as well as the acquisition and reconstruction parameters on explicit CT radiomics features. However, COVID-19 diagnosis can result in considering infected pulmonary lesions in both lungs (right and left), reconstruction parameters have the potential to enhance the model performance [12, 32, 44]. Future work should include different types of data (clinical and radiological) as prognostic indicators to develop a comprehensive model with a large sample size using multi-centre data to maximize the model performance.

## 5. Conclusion

CT radiomics features can be utilized towards derivation of biomarkers for COVID-19 and pneumonia diagnosis and severity prediction in a single 2-phase ML-based model. In terms of accuracy, the ML-based model demonstrated high performance in classifying COVID-19 and pneumonia (98%) cases using CT radiomics. In the second phase, an accuracy of 97% was achieved in classifying mild, moderate, and severe diseases. Our proposed, validated 2-phase model demonstrated great potential in assessing COVID-19 CT images towards rapid, reliable assessment and effective management of patients.

## Data Availability

All data produced in the present study are available upon reasonable request to the authors.

## Declaration of competing interest

The authors declare no conflicts of interest.

## Acknowledgements

This work was supported by the Omani Research Council Grant, grant number RC/COVID-MED/RADI/20/01.

## Notes

### Competing Interest Statement

The authors have declared no competing interest.

### Author Declarations

The SQUH and ROYH medical research ethical committees both approved this retrospective study (MREC#1254-REF. NO. SQU-EC/121/20)

